# A Systematic Review of Testing and Evaluation of Healthcare Applications of Large Language Models (LLMs)

**DOI:** 10.1101/2024.04.15.24305869

**Authors:** Suhana Bedi, Yutong Liu, Lucy Orr-Ewing, Dev Dash, Sanmi Koyejo, Alison Callahan, Jason A. Fries, Michael Wornow, Akshay Swaminathan, Lisa Soleymani Lehmann, Hyo Jung Hong, Mehr Kashyap, Akash R. Chaurasia, Nirav R. Shah, Karandeep Singh, Troy Tazbaz, Arnold Milstein, Michael A. Pfeffer, Nigam H. Shah

**Affiliations:** Stanford University; Stanford; Harvard University; University of California San Diego; US Food and Drug Administration

**Keywords:** Large Language Models, Generative Artificial Intelligence, Healthcare, Dimensions of Evaluation, Evaluation Metrics

## Abstract

1

**Importance:** Large Language Models (LLMs) can assist in a wide range of healthcare-related activities. Current approaches to evaluating LLMs make it difficult to identify the most impactful LLM application areas.

**Objective:** To summarize the current evaluation of LLMs in healthcare in terms of 5 components: evaluation data type, healthcare task, Natural Language Processing (NLP)/Natural Language Understanding (NLU) task, dimension of evaluation, and medical specialty.

**Data Sources:** A systematic search of PubMed and Web of Science was performed for studies published between 01-01-2022 and 02-19-2024.

**Study Selection:** Studies evaluating one or more LLMs in healthcare.

**Data Extraction and Synthesis:** Three independent reviewers categorized 519 studies in terms of data used in the evaluation, the healthcare tasks (the what) and the NLP/NLU tasks (the how) examined, the dimension(s) of evaluation, and the medical specialty studied.

**Results:** Only 5% of reviewed studies utilized real patient care data for LLM evaluation. The most popular healthcare tasks were assessing medical knowledge (e.g. answering medical licensing exam questions, 44.5%), followed by making diagnoses (19.5%), and educating patients (17.7%). Administrative tasks such as assigning provider billing codes (0.2%), writing prescriptions (0.2%), generating clinical referrals (0.6%) and clinical notetaking (0.8%) were less studied. For NLP/NLU tasks, the vast majority of studies examined question answering (84.2%). Other tasks such as summarization (8.9%), conversational dialogue (3.3%), and translation (3.1%) were infrequent. Almost all studies (95.4%) used accuracy as the primary dimension of evaluation; fairness, bias and toxicity (15.8%), robustness (14.8%), deployment considerations (4.6%), and calibration and uncertainty (1.2%) were infrequently measured. Finally, in terms of medical specialty area, most studies were in internal medicine (42%), surgery (11.4%) and ophthalmology (6.9%), with nuclear medicine (0.6%), physical medicine (0.4%) and medical genetics (0.2%) being the least represented.

**Conclusions and Relevance:** Existing evaluations of LLMs mostly focused on accuracy of question answering for medical exams, without consideration of real patient care data. Dimensions like fairness, bias and toxicity, robustness, and deployment considerations received limited attention. To draw meaningful conclusions and improve LLM adoption, future studies need to establish a standardized set of LLM applications and evaluation dimensions, perform evaluations using data from routine care, and broaden testing to include administrative tasks as well as multiple medical specialties.

**Key Points:** *Question:* How are healthcare applications of large language models (LLMs) currently evaluated?

*Findings:* Studies rarely used real patient care data for LLM evaluation. Administrative tasks such as generating provider billing codes and writing prescriptions were understudied. Natural Language Processing (NLP)/Natural Language Understanding (NLU) tasks like summarization, conversational dialogue, and translation were infrequently explored. Accuracy was the predominant dimension of evaluation, while fairness, bias and toxicity assessments were neglected. Evaluations in specialized fields, such as nuclear medicine and medical genetics were rare.

*Meaning:* Current LLM assessments in healthcare remain shallow and fragmented. To draw concrete insights on their performance, evaluations need to use real patient care data across a broad range of healthcare and NLP/NLU tasks and medical specialties with standardized dimensions of evaluation.

## 2. Introduction

The adoption of Artificial Intelligence (AI) in healthcare is rising, catalyzed by the emergence of Large Language Models (LLMs) like OpenAI’s ChatGPT ^1^ ^2^ ^3^ ^4^. Unlike predictive AI, generative AI produces original content such as sound, image, and text^5^. Within the realm of generative AI, LLMs produce structured, coherent prose in response to text inputs, with broad application in health system operations ^6^. Prominent applications such as facilitating clinical note-taking have already been implemented by several health systems in the U.S., and there is excitement in the medical community for improving healthcare efficiency, quality, and patient outcomes ^7^ ^8^. A recent report estimates that LLMs could unlock a substantial portion of the $1 trillion in untapped healthcare efficiency improvements, including an estimated savings ranging from 5 to 10 percent of US healthcare spending or approximately $200 billion to $360 billion annually based on 2019 figures ^9^ ^10^.

New and revolutionary technologies are often met with excitement about their many potential uses, leading to widespread and often unfocussed experimentation across different healthcare applications. Thus, as expected, the performance of LLMs in real-world healthcare settings too, remains inconsistently conducted and evaluated ^11^ ^12^. For instance, Cadamuro et al. assessed ChatGPT-4’s diagnostic ability by evaluating relevance, correctness, helpfulness, and safety, finding responses to be generally superficial and sometimes inaccurate, lacking in helpfulness and safety ^13^. In contrast, Pagano et al. also assessed diagnostic ability, but focused solely on correctness, concluding that ChatGPT-4 exhibited a high level of accuracy comparable to clinician responses ^14^.Thus, we hypothesize that the current evaluation landscape lacks the uniformity, thoroughness, and robustness necessary to effectively guide the deployment of LLMs in a real-world setting.

Our research aimed to assess the evaluation landscape of LLMs’ healthcare applications with the goal of accelerating their time to impact and successful integration. Through a systematic review of 519 studies, we comprehensively categorized how LLMs have been evaluated in healthcare along 5 axes: evaluation data type used, healthcare task, NLP/NLU task, dimension of evaluation, and medical specialty. To enable the categorization of the diverse range of applications and their evaluation setups, we used two categorization frameworks: the first describes healthcare applications of LLMs in terms of their constituent healthcare and NLP/NLU tasks, and the second describes dimensions of evaluation and associated metrics. These frameworks were then applied systematically to characterize the current state of evaluations to quantify the variability in LLM application evaluations and identify areas for further exploration.

Among the studies reviewed, there was no existing categorization framework that was consistently used. This lack of standardization required the creation of categorization framework, where we define each category as well as show illustrative examples of each. While such categorization can be a limitation, given that it provides a way to consistently discuss the testing and evaluations of LLM applications, it may have use beyond this review.

Our results show that evaluations of LLM applications in healthcare have been unevenly distributed both in terms of dimensions of evaluation used and in terms of medical specialty and application.

## 3. Methods

### 3.1 Design

A systematic review was conducted following Preferred Reporting Items for Systematic Reviews and Meta-Analyses (PRISMA) guidelines as shown in **Figure 1** ^15^.

**Figure 1:**
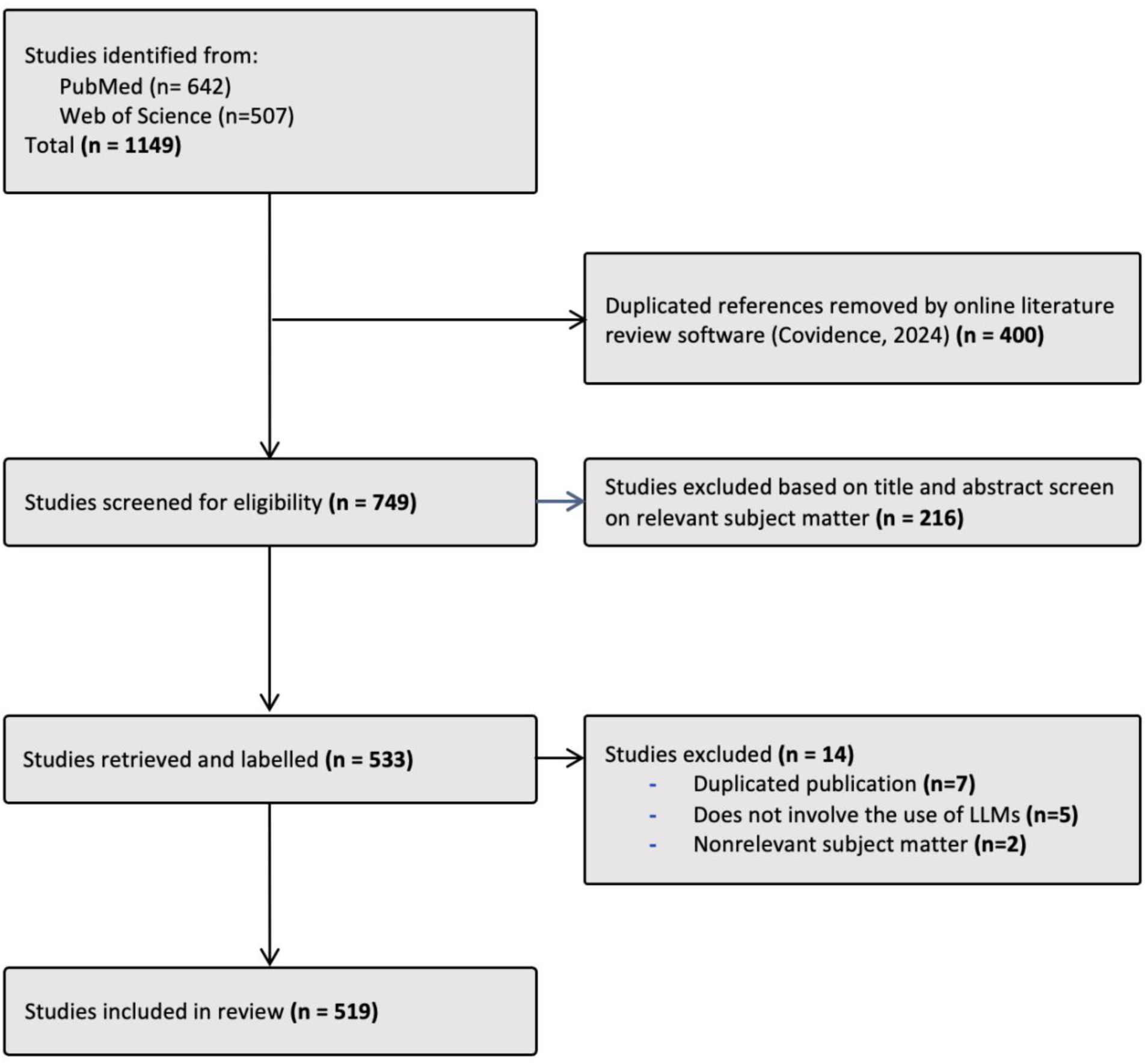
PRISMA Flow Diagram. This diagram shows the process of screening and selecting the categorized 519 studies.

### 3.2 Information sources

Peer-reviewed studies and preprints from January 1 2022, to February 19, 2024, were retrieved from PubMed and Web of Science databases, using specific keywords as detailed in **Supplement 1.** Our search focused on titles and abstracts to identify studies on evaluation of LLMs’ healthcare applications. This two-year period aimed to capture publications evaluating LLM healthcare applications since the public launch of ChatGPT in November 2022. Given our hypothesis that the current landscape lacks the necessary elements needed to truly assess LLM performance in healthcare, we included a broad spectrum of studies. Citations were imported into EndNote 21 (Clarivate) for analysis.

### 3.3 Categorization framework

Each study was categorized by evaluation data type, healthcare task, NLP/NLU task, dimension of evaluation, and medical specialty. Healthcare task categories were developed using publicly available healthcare task and competency lists and were refined by consulting board-certified MDs ^16^ ^17^ as outlined in **Table 1**. NLP/NLU categories and dimension of evaluations were developed using the Holistic Evaluation of Language Models (HELM) and Hugging Face frameworks ^18^ ^19^ as shown in **Tables 2** and **3**. Medical specialties were adapted from Accreditation Council for Graduate Medical Education (ACGME) residency programs. ^20^

**Table 1:** Healthcare task definitions and examples. This table lists the range of healthcare tasks that the 519 studies were categorized into, with definition and example for each task category.

| Healthcare Tasks | Definition | Example |
| --- | --- | --- |
| **Enhancing medical knowledge** | The process of enhancing the skills, knowledge, and capabilities of healthcare professionals to meet the evolving needs of healthcare delivery. | Measuring the performance of GPT on Neurosurgery Written Board examinations (Ali et al)<sup>21</sup> |
| **Making diagnoses** | The process of identifying the nature or cause of a disease or condition through the examination of symptoms, medical history, and diagnostic tests. | Comparing the performance of GPT and Physicians for diagnostic accuracy (Fraser et al)<sup>22</sup> |
| **Educating patients** | Providing patients with information and resources to help them understand their health conditions, treatment options etc. for more informed decision-making around their care. | Using GPT for Patient Information in periodontology (Babayigit et al)<sup>23</sup> |
| **Making treatment recommendations** | The process of providing treatment recommendations for patients to manage or cure their health conditions. | Using GPT for therapy recommendations in mental health (Patient Information in Periodontology (Wilhelm et al)<sup>24</sup> |
| **Communicating with patients** | The exchange of information between healthcare providers and patients. This could be done via patient messaging platforms, or via Chatbots integrated into the provider workflow. | Using GPT to communicate with palliative care patients (Srivastava et al)<sup>25</sup> |
| **Care coordination and planning** | The process of organizing and integrating healthcare services to ensure that patients receive the right care at the right time, involving communication and collaboration | Measuring the reliability and quality of nursing care planning generated (Dağcı et al)<sup>26</sup> |
| **Triaging patients** | Clinical triage is the process of prioritizing patients based on the severity of their condition and the urgency of their need for care. | Measuring the accuracy of patient triage in parasitology examination (Huh)<sup>27</sup> |
| **Carrying out a literature review** | A literature review is a critical summary and evaluation of existing research or literature on a specific topic. | Examining the validity of ChatGPT in identifying relevant Nephrology literature (Suppadungsuk et al)<sup>28</sup> |
| **Synthesizing data for research** | Data synthesis refers to the process of combining and analyzing data from multiple sources to generate new insights, draw conclusions, or develop a comprehensive understanding of a topic. | Synthesizing radiologic data for effective clinical decision-making (Rao et al)<sup>29</sup> |
| **Generating clinical referrals** | A referral is an order that a medical provider places to send their patient to a specialized physician or department for further evaluation, diagnosis, or treatment. | Assistance in optimizing Emergency Department radiology referrals and imaging selection (Barash et al)<sup>30</sup> |
| **Generating medical reports** | An image-captioning task of producing a professional report according to input image data. | Assessing the feasibility and acceptability of ChatGPT generated radiology report summaries for cancer patients (Chung et al)<sup>31</sup> |
| **Managing clinical knowledge** | The process of ensuring clinical knowledge bases is correct, consistent, complete, and current. | Using GPT models for phenotype concept recognition (Groza et al)<sup>32</sup> |
| **Providing asynchronous care** | A proactive way to ensure that everyone assigned to a clinic is up to date on basic preventive care - like cancer screenings or immunizations - and that they receive extra help if they have lab numbers that are high. | Asynchronously answering patient questions pertaining to erectile dysfunction (Razdan et al)<sup>33</sup> |
| **Clinical note-taking** | The process of recording detailed information about a patient's health status, medical history, symptoms, physical examination findings, diagnostic test results, treatment plans, typically documented in the patient's EMR | Using GPT models for taking notes during primary care visits (Kassab et al)<sup>34</sup> |
| **Enhancing surgical operations** | The process of supporting healthcare professionals, such as surgical technologists, nurses, and other staff, during surgical procedures | Using GPT to pinpoint innovations for future advancements in general surgery (Lim et al)<sup>35</sup> |
| **Conducting medical research** | Medical research generation, including writing papers, refers to the process of conducting original research in medicine or healthcare and documenting the findings in academic papers. | Using GPT models for sentiment analysis of COVID-19 survey data (Lossio-Ventura et al)<sup>36</sup> |
| **Biomedical data mining** | The process of searching and extracting data regarding a patient's health | Using GPT models to mine and generate biomedical text (Chen et al)<sup>37</sup> |
| **Generating provider billing codes** | Medical billing is the process of submitting and following up on claims with health insurance companies to receive payment for healthcare services provided to patients. | Using GPT models to predict diagnosis-related group (DRG) codes for hospitalized patients (Wang et al)<sup>38</sup> |
| **Writing prescriptions** | The process by which a healthcare provider, typically a physician or other qualified medical professional, orders medications or treatments for a patient | Prescription of kidney stone prevention treatment (Alumtrakul et al)<sup>39</sup> |

### 3.4 Eligibility criteria and screening

Screening was conducted by SB, YL, and LOE using the Covidence software (Covidence, 2024) as outlined in **Figure 1**. Included studies used LLMs for healthcare tasks and evaluated their performance. Excluded articles were those focused on multimodal tasks or basic biological science research with LLMs.

### 3.5 Data extraction and labeling

We adopted a paired review approach, wherein each study was categorized into evaluation data type, healthcare tasks, NLP/NLU tasks, dimension(s) of evaluation, and medical specialty by at least one human reviewer (SB, YL, or LOE) and GPT-4, based on the title and abstract. Note that GPT-4 was used as a force multiplier while the final categories were assigned by the human reviewers. In instances of disagreements regarding category assignments, the methods sections of the studies were retrieved, and final categories were determined through reviewer consensus. The prompts given to GPT-4 can be found in **Supplement 2.**

Each study received one or more healthcare tasks, NLP/NLU task, and dimension of evaluation labels as appropriate, hence the percentages sum above 100% in **Table 4**. In addition, each study could be assigned more than one medical specialty based on the evaluation conducted.

## 4. Results

749 relevant studies were screened for eligibility. After applying the inclusion and exclusion criteria described in **3.4**, 519 studies were included in the analysis using the frameworks developed by the authors.

### 4.1 Categorization framework for healthcare tasks, NLP/NLU tasks and dimensions of evaluation

We deconstructed each healthcare application of an LLM into its constituent healthcare task (**Table 1**), i.e. the clinical and non-clinical task it is used for (the “what”), and the NLP/NLU task (**Table 2**), i.e. the language processing task being performed (the “how”). Examples of a healthcare task are diagnosing a patient’s disease, recommending a treatment for osteoarthritis. Examples of the language-processing job to be accomplished – which is not necessarily specific to the medical domain are summarizing the impression section of a radiology report, answering questions about the symptoms of type 2 diabetes etc.

**Table 2:** Definition of NLP/NLU tasks. This table lists the range of NLP/NLU tasks that the 519 studies were categorized into, with definition and examples for each task category.

| **NLP/NLU Task** | **Definition** | **Examples** |
| --- | --- | --- |
| **Summarization** | For a clinical document $D$ of length $L$, generate a concise summary such that length of the summary $I \ll L$. | "Summarize the impression section of a radiology report" |
| **Question Answering** | For a clinical question $Q$, with or without reference to a context $T$, generate a response $R$. | - "What are the symptoms of Type 2 diabetes?"<br>- "What is the recommended dosage of ibuprofen for a 40 year-old male with mild fever?" |
| **Information Extraction** | For a clinical document $D$, extract structured information with semantic labels $s_1, ..., s_n$. | "Extract the mentions of adverse drug events, disease exacerbations and surgical interventions from a patient's history." |
| **Text Classification** | For a clinical document $D$ of length $L$, assign a label or class $P$ | "Categorize clinical notes into classes such as 'diagnosis', 'treatment' or 'prognosis'" |
| **Translation** | For a clinical document $D$ in language $M$, generate another document $D'$ in language $M'$ where $D = D'$ | "Translate a patient's old lab test results from Spanish to English" |
| **Conversational Dialogue** | For a history of chat messages $m_1, ..., m_n$ generate the next response $m_{n+1}$ | "Using a patient's history of chat messages, help them reschedule their appointment" |

An example of how healthcare tasks and NLP/NLU combine for a healthcare application of LLM is how Gan et al. evaluated LLM performance for mass-casualty triaging ^40^. The healthcare task (the “what”) is triaging patients while the NLP/NLU tasks (the “how”) are information extraction (extracting detailed patient information from the triage questionnaire scenarios, including age, symptoms, and vital signs), text classification (classifying the triage questionnaire scenarios into different triage levels), and question answering (generating final decision responses to the triage questionnaire).

We initially compiled a list of healthcare tasks using publicly available resources ^16^ ^17^. Subsequently, through consultation with three board-certified MDs, we refined the list through iterative discussions to establish the final categories for classification, as outlined in **Table 1**. To compile a list of common NLP/NLU tasks, we referred to sources such as the Holistic Evaluation of Language Models (HELM) study and the Hugging Face task framework to derive 6 categories: 1) Summarization, 2) Question answering, 3) Information extraction, 4) Text classification (such as clinical notes, research articles, and documents), 5) Translation, and 6) Conversational dialogue (**Table 2**) ^18^ ^19^.

We categorized the most common dimensions of evaluation used in the reviewed studies based on the list outlined in **Table 3**. These dimensions include: 1) Accuracy, 2) Calibration and uncertainty, 3) Robustness, 4) Factuality, 5) Comprehensiveness, 6) Fairness, bias, and toxicity, and 7) Deployment considerations. Fairness, bias, and toxicity were grouped together for ease of analysis, due to their infrequent occurrence in the reviewed studies, and relevance to ethical evaluation of LLMs. Additionally, we compiled common metrics for each dimension (**eTable 1**) to serve as a starting framework for researchers designing studies to assess LLM performance in healthcare applications.

**Table 3:** Dimensions of evaluation for LLM response. This table lists the range of dimensions of evaluations that the 519 studies were categorized into, with definitions, metrics and reviewer-generated example responses where each dimension is evaluated for a simple input question, “What are the symptoms of Type 2 diabetes?”

| Dimension of Evaluation | Definition | Metric Examples | Illustrative response demonstrating each dimension of evaluation |
| --- | --- | --- | --- |
| **Accuracy** | Measures how close the LLM output is to the true or expected answer | Human evaluated correctness, ROUGE, MEDCON | **Correct Response** - "Common symptoms of Type 2 Diabetes include frequent urination, increased thirst, unexplained weight loss, fatigue, and blurred vision." |
| **Calibration and Uncertainty** | Measures how uncertain or underconfident an LLM is about its output for a specific task | Human evaluated uncertainty, calibration error, Platt scaled calibration slope | **Response with an uncertainty estimate** - "As per my knowledge, the most common symptoms of Type 2 Diabetes are frequent urination, increased thirst, and unexplained weight loss, however, my information might be outdated, so I would put a confidence score 0.3 for my response and I would recommend contacting a healthcare provider for a more accurate and certain response." |
| **Robustness** | Measures the LLM's resilience against adversarial attacks and perturbations like typos. | Human evaluated robustness, exact match on LLM input with intentional typos, F1 on LLM input with intentional use of word synonyms | **Variation 1**: "What are the signs of Type 2 Diabetes?"<br>**Robust Response (Synonym)**: "Signs of Type 2 Diabetes include frequent urination, increased thirst, unexplained weight loss, fatigue, and blurred vision."<br>**Variation 2 (Typo)**: "Symptom of Tpye 2 Diabetes?"<br>**Robust Response**: "Symptoms of Type 2 Diabetes include frequent urination, increased thirst, unexplained weight loss, fatigue, and blurred vision." |
| **Factuality** | Measures how an LLM's output for a specific task originates from a verifiable and citable source. It is **important** to note that it is possible for a response to be accurate but factually incorrect if it originates from a hallucinated citation | Human evaluated factual consistency, citation recall, citation precision | **Factual Response**: "Symptoms of Type 2 Diabetes are often related to insulin resistance and include frequent urination, increased thirst, unexplained weight loss, fatigue, and blurred vision. Here is a reference to the link I referred to in crafting this response - https://www.niddk.nih.gov/health-information/diabetes/overview/what-is-diabetes/type-1-diabetes" |
| **Comprehensiveness** | Measures how well an LLM's output coherently and concisely addresses | Human evaluated comprehensiveness, fluency, UniEval relevance | **Comprehensive Response**: "Symptoms of Type 2 Diabetes include frequent urination, increased thirst, unexplained weight loss, fatigue, blurred vision, slow |
|  | all aspects of the task and reference provided |  | wound healing, and tingling or numbness in the hands or feet. " |
| **Fairness, bias and toxicity** | Measures whether an LLM's output is equitable, impartial, and free from harmful stereotypes or biases, ensuring it does not perpetuate injustice or toxicity across diverse groups | Human evaluated toxicity, counterfactual fairness, performance disparities across race | **Unbiased Response:** "Symptoms of Type 2 Diabetes can vary, and it's important to seek medical advice for proper diagnosis. Common symptoms include frequent urination, increased thirst, unexplained weight loss, fatigue, and blurred vision."<br>**Biased Response:** "Type 2 Diabetes symptoms are often seen in individuals with poor lifestyle choices." |
| **Deployment considerations** | Measures the technical and parametric details of an LLM to generate a desired output | Cost, latency, inference runtime | **Response with runtime:** "The model provides information about Type 2 Diabetes symptoms in less than 0.5 second, ensuring quick access to essential health information." |

### 4.2 Distribution of studies based on evaluation data type

Among the reviewed studies, 5% evaluated and tested LLMs using real patient care data, while the remaining relied on data such as medical examination questions, clinician-designed vignettes or Subject Matter Expert (SME) generated questions.

### 4.3 Categorizing articles based on healthcare tasks and NLP/NLU tasks

The studies we examined had a predominant focus on evaluating LLMs for their medical knowledge (**Table 4)**, primarily through assessments such as the USMLE. This trend assumes that because we assess medical professionals’ readiness for entering clinical practice through board-style examinations, mirroring this type of evaluation for LLMs is adequate to certify their fitness-for-use. Making diagnoses, educating patients and making treatment recommendations were the other common healthcare tasks studied. While these tasks represent critical aspects of healthcare delivery, validating the utility of LLMs in supporting them requires assessment with real patient care data. The limited examination of administrative tasks like assigning provider billing codes, writing prescriptions, generating clinical referrals, and clinical notetaking suggests a gap in studying LLMs’ use for high-value, immediately impactful administrative tasks. These tasks are often labor intensive, presenting a ripe opportunity for testing LLMs to enhance efficiency in these areas ^41^.

**Table 4:** Frequency of publications examining each dimension of evaluation across healthcare and NLP/NLU task categories. The first column lists healthcare tasks followed by NLP/NLU tasks (separated by a double line); the first row lists the dimensions of evaluation used in each study examined. The percentages in the last row are the percentage of studies in which a specific dimension was evaluated and the percentages on the last column indicate the percentage of studies in which a specific healthcare task or NLP/NLU task was evaluated.

|  | Accuracy | Comprehensiveness | Factuality | Robustness | Fairness, Bias and Toxicity evaluation | Deployment Metrics | Calibration and Uncertainty | TOTAL NUMBER OF PAPERS |  |
| --- | --- | --- | --- | --- | --- | --- | --- | --- | --- |
|  |  |  |  |  |  |  |  | Number | % |
| **Healthcare Task** |  |  |  |  |  |  |  |  |  |
| Enhancing medical knowledge | 222 | 91 | 44 | 33 | 16 | 10 | 3 | 231 | 44.5% |
| Making Diagnoses | 100 | 38 | 11 | 11 | 14 | 4 | 0 | 101 | 19.5% |
| Educating patients | 88 | 68 | 32 | 21 | 18 | 3 | 2 | 92 | 17.7% |
| Making treatment recommendations | 47 | 22 | 9 | 8 | 3 | 1 | 0 | 48 | 9.2% |
| Communicating with patients | 35 | 29 | 8 | 15 | 22 | 1 | 0 | 39 | 7.5% |
| Care coordination and planning | 36 | 24 | 4 | 5 | 7 | 1 | 0 | 39 | 7.5% |
| Triaging patients | 24 | 7 | 5 | 2 | 8 | 8 | 0 | 24 | 4.6% |
| Carrying out a literature review | 18 | 7 | 3 | 2 | 2 | 2 | 0 | 18 | 3.5% |
| Synthesizing data for research | 16 | 7 | 2 | 3 | 2 | 2 | 0 | 17 | 3.3% |
| Generating medical reports | 8 | 8 | 2 | 0 | 3 | 0 | 0 | 9 | 1.7% |
| Conducting medical research | 8 | 7 | 3 | 3 | 3 | 1 | 0 | 9 | 1.7% |
| Providing asynchronous care | 8 | 5 | 3 | 3 | 1 | 1 | 0 | 8 | 1.5% |
| Managing clinical knowledge | 5 | 5 | 1 | 1 | 0 | 0 | 0 | 6 | 1.2% |
| Clinical note-taking | 6 | 2 | 1 | 1 | 0 | 0 | 1 | 4 | 0.8% |
| Generating clinical referrals | 3 | 0 | 0 | 0 | 0 | 1 | 0 | 3 | 0.6% |
| Enhancing surgical operations | 3 | 3 | 1 | 1 | 0 | 0 | 0 | 3 | 0.6% |
| Biomedical data mining | 2 | 0 | 0 | 2 | 1 | 2 | 0 | 2 | 0.4% |
| Generating provider billing codes | 1 | 0 | 0 | 0 | 0 | 0 | 0 | 1 | 0.2% |
| Writing prescriptions | 1 | 0 | 0 | 0 | 0 | 0 | 0 | 1 | 0.2% |
| **NLP/NLU Task** |  |  |  |  |  |  |  |  |  |
| Question Answering | 398 | 194 | 71 | 61 | 54 | 14 | 5 | 437 | 84.2% |
| Text Classification | 29 | 10 | 6 | 5 | 10 | 2 | 0 | 145 | 27.9% |
| Information Extraction | 29 | 12 | 8 | 5 | 4 | 6 | 0 | 128 | 24.7% |
| Summarization | 29 | 21 | 7 | 3 | 8 | 0 | 1 | 46 | 8.9% |
| Conversational Dialogue | 6 | 6 | 1 | 1 | 5 | 1 | 0 | 17 | 3.3% |
| Translation | 5 | 1 | 2 | 2 | 1 | 1 | 0 | 16 | 3.1% |
| **TOTAL NUMBER OF PAPERS** | 495 | 244 | 95 | 77 | 82 | 24 | 6 |  |  |
| % | 95.4% | 47.0% | 18.3% | 14.8% | 15.8% | 4.6% | 1.2% |  |  |

Among the NLP/NLU tasks, most studies evaluated LLM performance through question answering tasks. These tasks ranged from addressing generic inquiries about symptoms and treatments to tackling board-style questions featuring clinical vignettes. Approximately a quarter of the studies focused on text classification and information extraction tasks. Tasks such as summarization, conversational dialogue, and translation remained underexplored. This gap is significant because condensing patient records into concise summaries, translating medical content into simpler languages or the patient’s native language, and facilitating conversations through chatbots are often touted benefits of using LLMs and could substantially alleviate physician burden.

### 4.4 Categorizing articles based on the dimensions of evaluation

As seen in **Table 4**, accuracy and comprehensiveness were overwhelmingly the top two most examined dimensions, whereas factuality, fairness, bias, and toxicity, robustness, deployment considerations, and calibration and uncertainty were infrequently assessed. This suggests a potential gap in assessing the broader capabilities and suitability of LLMs for real-world deployment. While accuracy and comprehensiveness are crucial for ensuring the reliability and effectiveness of LLMs in healthcare tasks, dimensions like fairness, bias, and toxicity are equally vital for addressing ethical concerns and ensuring equitable outcomes. Similarly, robustness and deployment considerations are essential for assessing the sustainability of integrating LLMs into healthcare systems. The limited assessment of calibration and uncertainty raises questions about the extent to which researchers are addressing the need for LLMs to provide uncertainty quantifications, particularly in healthcare scenarios.

### 4.5 Distribution of studies by medical specialty

We categorized studies according to the Accreditation Council for Graduate Medical Education (ACGME) residency programs, augmented to include additional categories to capture studies investigating applications in dental specialties, treatment of genetic disorders and generic healthcare applications ^20^. Notably, over a fifth of the studies were categorized as generic, indicating a significant focus on healthcare applications that are relevant to many specialties, rather than a specific specialty. Among the specialties, internal medicine, surgery, and ophthalmology were the top specialties. Nuclear medicine, physical medicine, and medical genetics were the least prevalent specialties in studies, accounting for 12 studies in total. The exact percentage of studies in different specialties are outlined in **eTable 2**. The distribution of studies across specialties underscores the potential for LLMs to contribute to a wide range of medical specialties, but also signals opportunities for further exploration within less represented areas such as nuclear medicine, physical medicine, and medical genetics.

## 5. Discussion

Our systematic review of 519 studies summarizes existing evaluations of LLMs across medical specialties, the underlying healthcare task, NLP/NLU task, and dimension of evaluation. Doing such categorization captures the heterogeneity of current LLM applications in healthcare while providing a concrete way to discuss their testing and evaluation in a consistent manner. While our categorization we created may be viewed as a limitation, given the precise definitions and illustrative examples of each category, it may have utility beyond this review.

Overall, we identified six limitations in the current efforts and suggest how to address them in future. Our findings call for an urgent need to develop consensus-driven guidance for evaluating LLMs in medicine, in a manner similar to the creation of the blueprint for trustworthy AI by The Coalition for Health AI for traditional AI models^42^.

### The need for evaluations based on real patient care data

One striking finding is that only 5% of the studies used real patient care data for evaluation, with most studies using a mix of medical exam questions, patient vignettes and subject matter expert generated questions ^43^ ^14^ ^44^. Shah et al noted that testing LLMs with hypothetical medical questions is like assessing a car’s performance with multiple-choice questions before certifying it for road use ^11^. Real patient care data encompasses the complexities of clinical practice, providing a more thorough evaluation of LLM performance that will closely mirror real-world performance ^14^ ^45^ ^46^ ^6^.

Real-world LLM evaluations provide valuable insights that may be overlooked in simulations or synthetic environments. For instance, while LLMs have been touted for potentially saving time and enhancing clinician experience, Garcia et al. found that the mean utilization rate for drafting patient messaging responses in an EHR system was only 20%, resulting in a reduction in burnout score but no time savings ^47^.

Given the importance of using real patient care data, systems need to be created to ensure their use in evaluating LLMs’ healthcare applications. The Office of the National Coordinator for Health Information Technology (ONC) recently passed HT-1, the first federal regulation to set specific reporting requirements for developers of AI tools through their ‘model report cards’^48^. ONC and other regulators should look to embed a mandate for the use of patient care data in the evaluation process of LLM tools into its requirements.

### The need to standardize the task formulations and dimensions of evaluation

There is a lack of consensus on which dimensions of evaluation to examine for a given healthcare task or NLP/NLU task. For instance, for a medical education task, Ali et al. tested the performance of GPT-4 on a written board examination focusing on output accuracy as the sole dimension ^21^. Another study tested the performance of ChatGPT on the USMLE, focusing on output accuracy, factuality and comprehensiveness as primary dimensions of evaluation ^49^.

To address this challenge, we need to establish shared definitions of tasks and corresponding dimensions of evaluation. Similar to how efforts such as Holistic Evaluation of Language Models (HELM) define the dimensions of evaluation of an LLM that matter in general, a framework specific for healthcare is necessary to define the core dimensions of evaluation to be assessed across studies. Doing so enables better comparisons and cumulative learning from which reliable conclusions can be drawn for future technical work and policy guidance.

### Prioritize immediately impactful, administrative applications

Current research predominantly focuses on medical knowledge tasks, such as answering medical exam questions (44.5%), or complex healthcare tasks, as well as making diagnoses (19.5%) and making treatment recommendations (9.2%). However, there are many administrative tasks in healthcare that are often labor-intensive, requiring manual input and contributing to physician burnout ^41^. Particularly, areas such as assigning provider billing codes (1 study), writing prescriptions (1 study), generating clinical referrals (3 studies), and clinical note-taking (4 studies); all of which remain under-researched and could greatly benefit from a systematic evaluation of using LLMs for those tasks ^38^ ^39^ ^50^ ^51^.

### The need to bridge gaps in LLM utilization across clinical specialties

The substantial representation of generic healthcare applications, accounting for over a fifth of the studies, underscores the potential of LLMs in addressing needs applicable to many specialties, such as summarizing medical reports. In contrast, the scarcity of research in particular specialties like nuclear medicine (3 studies), physical medicine (2 studies), and medical genetics (1 study) suggests an untapped potential for using LLMs in these complex medical domains that often present intricate diagnostic challenges and demand personalized treatment approaches^52^ ^53^ ^54^ ^55^. The lack of LLM-focused studies in these areas may indicate the need for increased awareness, collaboration, or specialized adaptation of such models to suit the unique demands of these specialties.

### The need for a realistic accounting of financial impact

Generative AI is projected to create $200 billion to $360 billion in healthcare cost savings through productivity improvements ^10^. However, the implementation of these tools could pose a significant financial burden to health systems. In a recent review by Sahni and Carrus, defining the cost and benefit of deploying AI was highlighted as one of the greatest challenges ^56^. It is key for health systems to capture this, to accurately estimate and budget for increased implementation and computing costs ^57^.

Within this review, only one study conducted a financial impact or cost-effectiveness analysis. Rau et al. investigated the use of ChatGPT to develop personalized imaging, demonstrating “an average decision time of 5 minutes and a cost of €0.19 for all cases, compared to 50 minutes and €29.99 for radiologists” ^58^. However, this analysis was a parallel implementation of the LLM solution compared with the traditional radiologist approach, thus not providing a realistic assessment of the *added* value of LLM integration into existing clinical workflows and its corresponding financial impact.

While the dearth of real-world testing is understandable given the infancy of LLM applications in healthcare, it is imperative to establish realistic assessments of these tools before reallocating resources from other healthcare initiatives. Notably, such assessments should estimate the total cost of implementation, which includes not only the cost to run the model but also expenses associated with monitoring, maintenance, and any necessary infrastructure adjustments.

### The need to better define and quantify bias

Recent studies have highlighted a concerning trend of LLMs perpetuating race-based medicine in their responses ^59^. This phenomenon can be attributed to the tendency of LLMs to reproduce information from their training data, which may contain human biases ^60^.To improve our methods for evaluating and quantifying bias, we need to first collectively establish what it means to be unbiased.

While efforts to assess racial and ethical biases exist, only 15.8% of studies have conducted *any* evaluation that delves into how factors such as race, gender, or age impact bias in the model’s output ^61^ ^62^ ^63^. Future research should place greater emphasis on such evaluations, particularly as policymakers develop best practices and guidance for model assurance. Mandating these evaluations as part of a “model report card” could be a proactive step towards mitigating harmful biases perpetuated by LLMs ^64^.

### The need to publicly report failure modes

The analysis of failure modes has long been regarded as fundamental in engineering and quality management, facilitating the identification, examination, and subsequent mitigation of failures^65^. The FDA has databases for adverse event reporting in pharmaceuticals and medical devices, but there is currently no analogous place for reporting failure modes for AI systems, let alone LLMs, in healthcare ^66^ ^67^.

In the ‘Conclusion’ sections of many studies, only a select few researched why the deployment of the LLM did not produce satisfactory results (e.g. ineffective prompt engineering) ^68^. A deeper examination of failure modes and why the exercise was deemed unsuccessful or inaccurate (e.g. the reference data was factually incorrect or outdated), is necessary to further improve the use of LLMs in healthcare settings.

## 6. Conclusion

The evaluation of LLMs lacks standardized task definitions and dimensions of evaluation. This systematic review underscores the need for evaluating LLMs using real patient care data, particularly on administrative healthcare tasks like generating provider billing codes, writing prescriptions, and clinical note-taking. It highlights the need to expand testing criteria beyond accuracy to include fairness, bias, toxicity, robustness, and deployment considerations across different medical specialties. Establishing shared task definitions and rigorous testing and evaluation standards are crucial for the safe integration of LLMs in healthcare. Realistic financial accounting and robust reporting of failures are essential to accurately assess their value and safety in clinical settings. Broadly, there is an urgent need to develop a nationwide consensus and guidance for evaluating LLMs in healthcare, so that we may realize the tremendous promise these groundbreaking technologies have to offer.

## Author contributions

SB, YL, LOE, NHS conceived of the study, defined the main outcomes and measures. SB, LOE, YL searched the literature to identify the publications to review and categorized the publications. SB, LOE, YL and NHS drafted the manuscript. SB designed the GPT-4 based screening strategy with input from DD. SB developed the NLP/NLU task and dimensions of evaluation framework. LOE developed the healthcare task framework. DD guided the creation and categorization of healthcare tasks. AC and AS guided the creation of NLP/NLU task categorization. JAF guided the creation of the dimensions of evaluation categorization, SK helped select HELM dimensions to reuse. MK refined the medical specialty categorization, and MW critiqued the review methodology and figure organization. LL and HH assessed the usefulness of the frameworks for other analyses. NRS guided LOE and YL on all aspects of performing systematic reviews. AM reviewed and edited the manuscript for framing the discussion. KS and TT assessed the relevance of the results for developing consensus LLM testing and evaluation guidance for CHAI. MAP critiqued the deployment concerns in health systems and reviewed the categories. All authors reviewed, edited and approved of the final manuscript.

## Data Availability

All data produced in the present study are available upon reasonable request to the authors

## Acknowledgements

We thank Nicholas Chedid for extensive guidance in the development of the healthcare task categorization.

**Supplement 1. Search terms for PubMed as of 02/19/2024**

((“Large Language Model” [Title/Abstract] OR “ChatGPT” [Title/Abstract] OR “Generative AI” [Title/Abstract]) AND (“Health” [Title/Abstract] OR “Medical” [Title/Abstract] OR “Clinical” [Title/Abstract] OR “Medicine” [Title/Abstract]) AND (“Test” [Title/Abstract] OR “Evaluate” [Title/Abstract] OR “Performance” [Title/Abstract] OR “Assess” [Title/Abstract]))

**Search terms for Web of science as of 02/19/2024**

(TS=(“Large Language Model” OR “ChatGPT” OR “Generative AI”)

AND

TS=(”Health” OR “Medical” OR “Clinical” OR “Medicine”)

AND

TS=(“Test” OR “Evaluate” OR “Performance” OR “Assess”))

**Supplement 2. Prompts used to extract and assign categories for human review**

**Prompt 1**

“You are assisting in a systematic review of large language models in healthcare. Summarize the {entity_type} mentioned in this research abstract in 25 words”

**Prompt 2**

**“**Using the generated summaries, identify and categorize the following text based on {entity_type}:\n\n{text}\n\nCategories:**”**

Where entity_type can be *NLP task*, *medical specialty* or *metric* and categories is the list of possible values for each entity_type to make categorization into, for the NLP task, metric and medical specialty.

---

**eTable 1.**
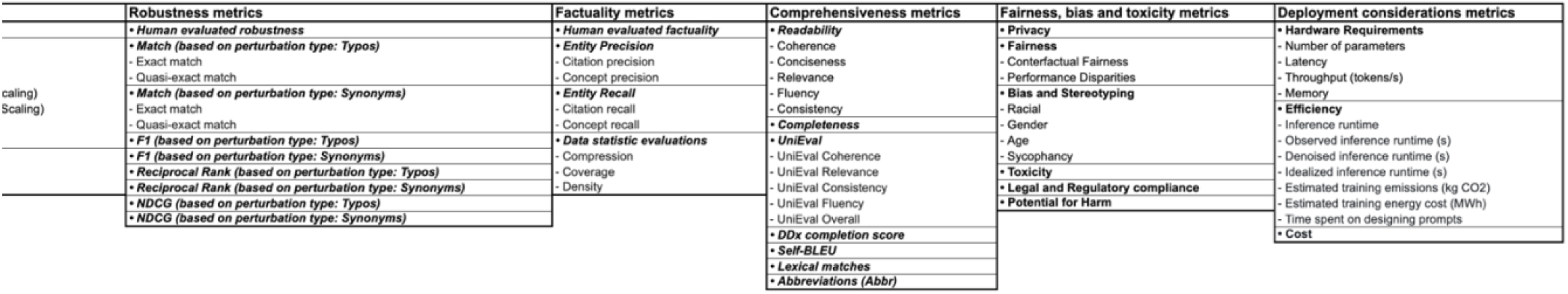
Examples of metrics for each dimension of evaluation. The first row represents the names of the dimensions of evaluation in our designed framework. Under each dimension there are metrics. The bold italicized cells represent metric subclasses for each dimension and regular font cells under each subclass represent the metrics.

**eTable 2.**
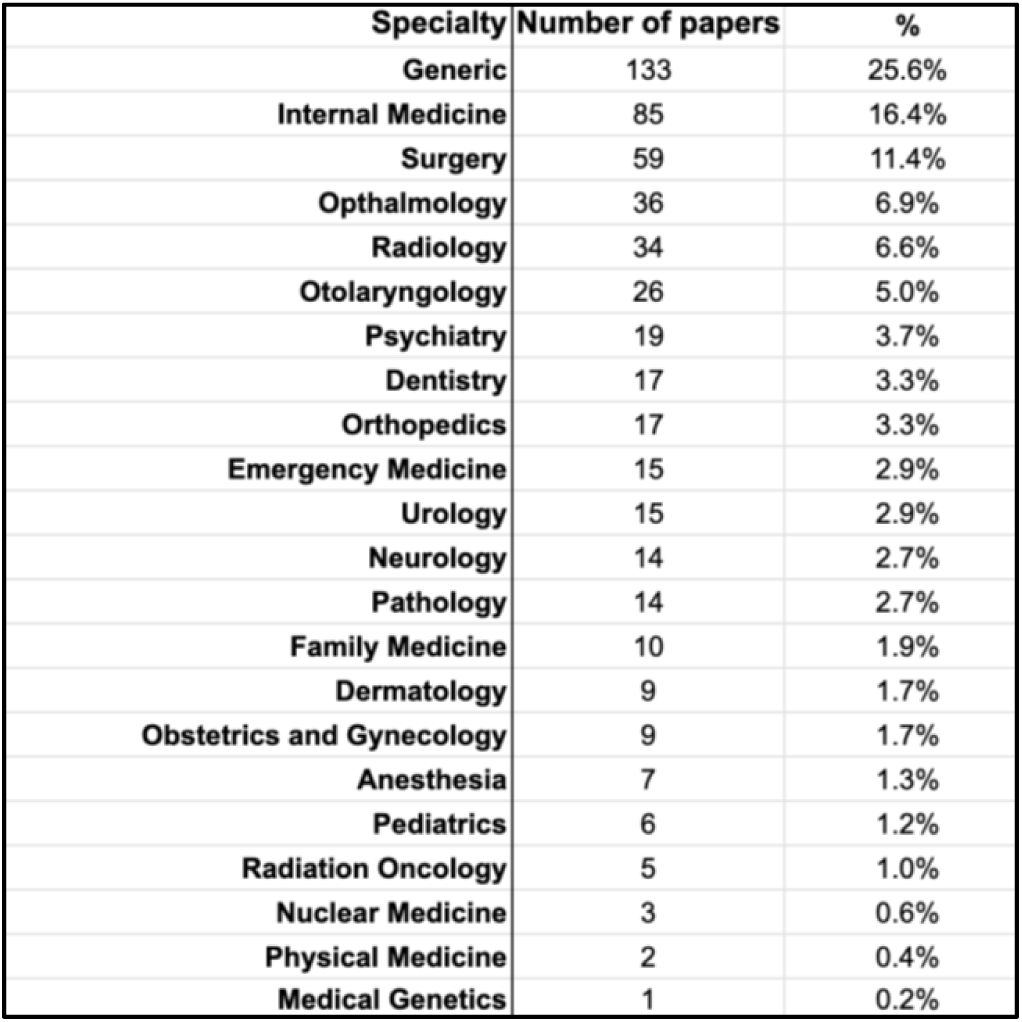
Frequency of publications by medical specialty. This table shows the different medical specialties of the 519 studies, along with three additional categories: Generic, Dentistry, and Medical Genetics

| Specialty | Number of papers | % |
| --- | --- | --- |
| Generic | 133 | 25.6% |
| Internal Medicine | 85 | 16.4% |
| Surgery | 59 | 11.4% |
| Ophthalmology | 36 | 6.9% |
| Radiology | 34 | 6.6% |
| Otolaryngology | 26 | 5.0% |
| Psychiatry | 19 | 3.7% |
| Dentistry | 17 | 3.3% |
| Orthopedics | 17 | 3.3% |
| Emergency Medicine | 15 | 2.9% |
| Urology | 15 | 2.9% |
| Neurology | 14 | 2.7% |
| Pathology | 14 | 2.7% |
| Family Medicine | 10 | 1.9% |
| Dermatology | 9 | 1.7% |
| Obstetrics and Gynecology | 9 | 1.7% |
| Anesthesia | 7 | 1.3% |
| Pediatrics | 6 | 1.2% |
| Radiation Oncology | 5 | 1.0% |
| Nuclear Medicine | 3 | 0.6% |
| Physical Medicine | 2 | 0.4% |
| Medical Genetics | 1 | 0.2% |

## Notes

### Competing Interest Statement

The authors have declared no competing interest.

### Funding Statement

This study did not receive any funding

### Summary of Updates

This version of the manuscript has been revised to display table 4 which was missing from the previous version.

